# Inequalities in air pollution exposure are increasing in the United States

**DOI:** 10.1101/2020.07.13.20152942

**Authors:** Abdulrahman Jbaily, Xiaodan Zhou, Jie Liu, Ting-Hwan Lee, Stéphane Verguet, Francesca Dominici

## Abstract

Exposure to ambient air pollution contributes substantially to the global burden of disease, and in 2015, ambient exposure to PM_2.5_ (fine particles with a mass median aerodynamic diameter of less than 2.5 *μm*) was the fifth-ranking risk factor of mortality globally. We analyzed data from the US zip code tabulation areas (N=32047) for 2000-2016 and found strong evidence of inequalities in exposure to PM_2.5_ among both racial/ethnic and income groups. Most alarming, we found that these inequalities have been increasing over time. From 2010 to 2016 inequalities in the exposure to PM_2.5_ levels above 8 *μg/m*^3^ across racial/ethnic, and income groups increased by factors of 1.6 and 4.0 respectively. As shown in our powerful map visualizations, these results indicate that air pollution regulations must not only decrease PM_2.5_ concentration levels nationwide but also prioritize reducing environmental injustice across the US.

---

Strong associations exist between exposure to PM_2.5_ (fine particles with a mass median aerodynamic diameter of less than 2.5 *μm*) in the US and adverse health outcomes such as hospital admissions [1, 2, 3, 4] and mortality [5, 6, 7, 8, 9, 10]. It is well documented that minorities and people of low socioeconomic status in the US are at a higher risk of death from being exposed to PM_2.5_ [11, 12, 13, 14]. Although inequalities in air pollution exposure among racial/ethnic and socioeconomic groups in the US are known to exist [15, 16, 17, 18], it is unknown whether these inequalities are being addressed. Here we provide strong and concerning evidence that although air pollution in the US has decreased over the years, disparities among racial/ethnic populations and socioeconomic groups have increased. We found that Black populations and lower income groups are exposed to the highest levels of particulate matter across the study period and that inequalities in exposure to particulate matter among racial/ethnic communities have doubled between 2010 and 2016. Our findings have strong and time-sensitive implications on policies set by the US Environmental Protection Agency (EPA) to combat racial injustice and the higher death toll of COVID-19 among minorities.

## Disparities in air pollution exposure among racial*/*ethnic groups

The US EPA is required to reexamine its National Ambient Air Quality Standards (NAAQS) every five years. In 2012 the EPA set the NAAQS for PM_2.5_ to 12 *μg/m*^3^ [19]. On average across the US, we found that PM_2.5_ has decreased by 43% (Animation 1 in supplementary material; and Extended Data figure A.1). We also found that the percentage of the population exposed to PM_2.5_ levels higher than 12 *μg/m*^3^ decreased from 62.8% in 2000 to 0.23% in 2016 (map shown in Extended Data figure A.2).

For each racial/ethnic group (White, Black, Asian, and Hispanic)^1^, we construct a map that shows only zip code tabulation areas (ZCTAs) where the race/ethnicity is most present^2^. Figures 1a and 1b show the PM_2.5_ distribution on the constructed Black and White population maps for 2000 and 2016, respectively. We found that the Black ZCTAs (left map) are dominated by high PM_2.5_ concentrations relative to the White ZCTAs (right map) in both 2000 and 2016. Furthermore, we see a steeper decline in PM_2.5_ among the White ZCTAs as compared to the Black ZCTAs. Similar maps for the Asian and Hispanic groups are shown in Extended Data figure A.3 and Animation 2 in supplementary material.

**Figure 1:**
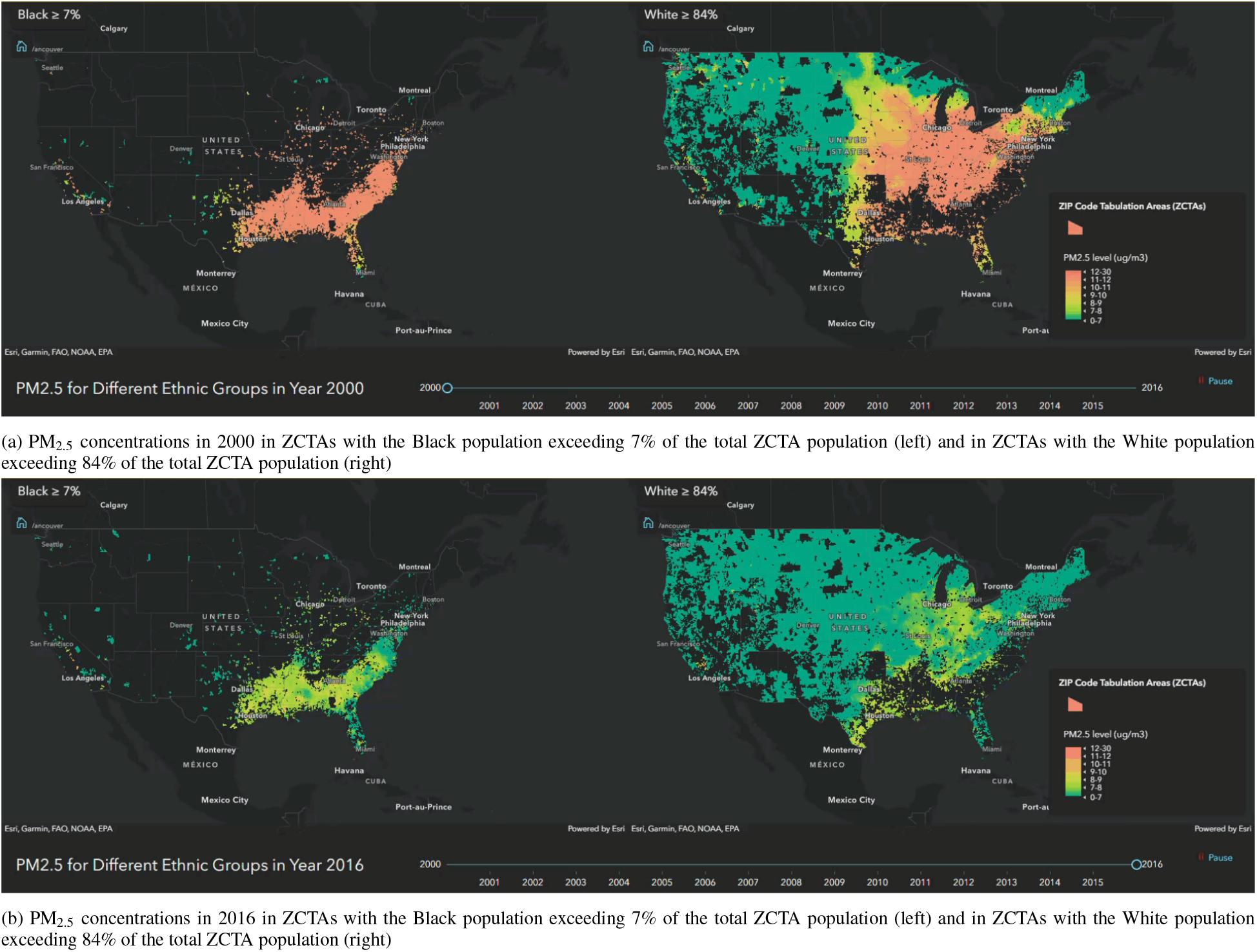
We use the White population fraction of the ZCTA population to compute the average White population fraction (aWpf) across all ZCTAs (≈ 84%). Similarly, we compute the average Black population fraction (aBpf) (≈ 7%). The maps in panel (a) show PM_2.5_ levels for the year 2000 in ZCTAs with a Black population fraction above aBpf (left) and in ZCTAs with a White population fraction aWpf (right). High PM_2.5_ concentrations exist in almost all ZCTAs with a Black population above aBpf, while a wide range of low and high PM_2.5_ concentrations exist in ZCTAs with a White population above aWpf in 2000. Panel (b) shows the same information for the year 2016. It can be visually seen that a larger decrease in PM_2.5_ concentrations from 2000 to 2016 occurred in ZCTAs corresponding to the White population (right map of panel b). Similar maps of the Hispanic and Asian populations for 2000 and 2016 are shown in Extended Data figures A.3a and A.3b and Animation 2 in supplementary material.

To quantify these differences in exposure, we compute the population-weighted average PM_2.5_ concentration for every racial/ethnic population (please see Methods) (Extended Data figure A.4a). For all years, we found that the Black population is exposed to higher levels of PM_2.5_ compared to other racial/ethnic groups. To visualize this finding for 2016, we illustrate how the population-weighted PM_2.5_ average concentration changes as ZCTAs become more populated by a certain race/ethnicity (Extended Data figure A.4b). We found that as the Black population increases in a ZCTA, the PM_2.5_ concentration consistently increases and a very steep incline is seen for ZCTAs with more than 85% of their population as Black. The trend of the Hispanic population follows that of the Black population except at population fractions above 85%. The opposite is seen for White populations; PM_2.5_ concentration decreases as density of the White population increases in ZCTAs; a steep decrease is shown for ZCTAs with a White population fraction exceeding 70%. Further, in ZCTAs where the population of Native Americans is at least 80%, the average PM_2.5_ concentration drops to below 4 *μg/m*^3^. When the Asian population size increases there is no notable change in the average PM_2.5_ concentration.

## Disparities in air pollution exposure among income groups

Disparities exist among distinct income groups, with low-income groups exposed to the highest PM_2.5_ concentrations. We assign all ZCTAs percentile ranks from 1 to 100 based on median household income and categorize them into ten income groups. We designate the lowest and highest three income groups as low-income and high-income respectively and then split the US map into two maps – ZCTAs defined as low- and high-income (please see Methods). We visualize the PM_2.5_ concentration distribution on the two maps for 2000 to 2016 (Animation 3 in supplementary material). The low-income map appears to be dominated by an overall higher concentration of PM_2.5_ as compared to the high-income map especially in recent years. We include snapshots of 2000 and 2016 (figure 2). In 2016 for example, ∼ 80% of ZCTAs of the high-income US map had PM_2.5_ concentrations lower than 8 *μg/m*^3^ compared to only ∼ 70% of the ZCTAs of the low-income map (figure 2b). We summarize the contents of the maps by computing the average PM_2.5_ concentration for the low- and high-income groups (Extended Data figure A.5); the PM_2.5_ concentration is consistently higher for the low-income group, except in 2001. Further, we isolate the effects of income on the disparities among the racial/ethnic groups in Extended Data figures A.6a and A.6b.

**Figure 2:**
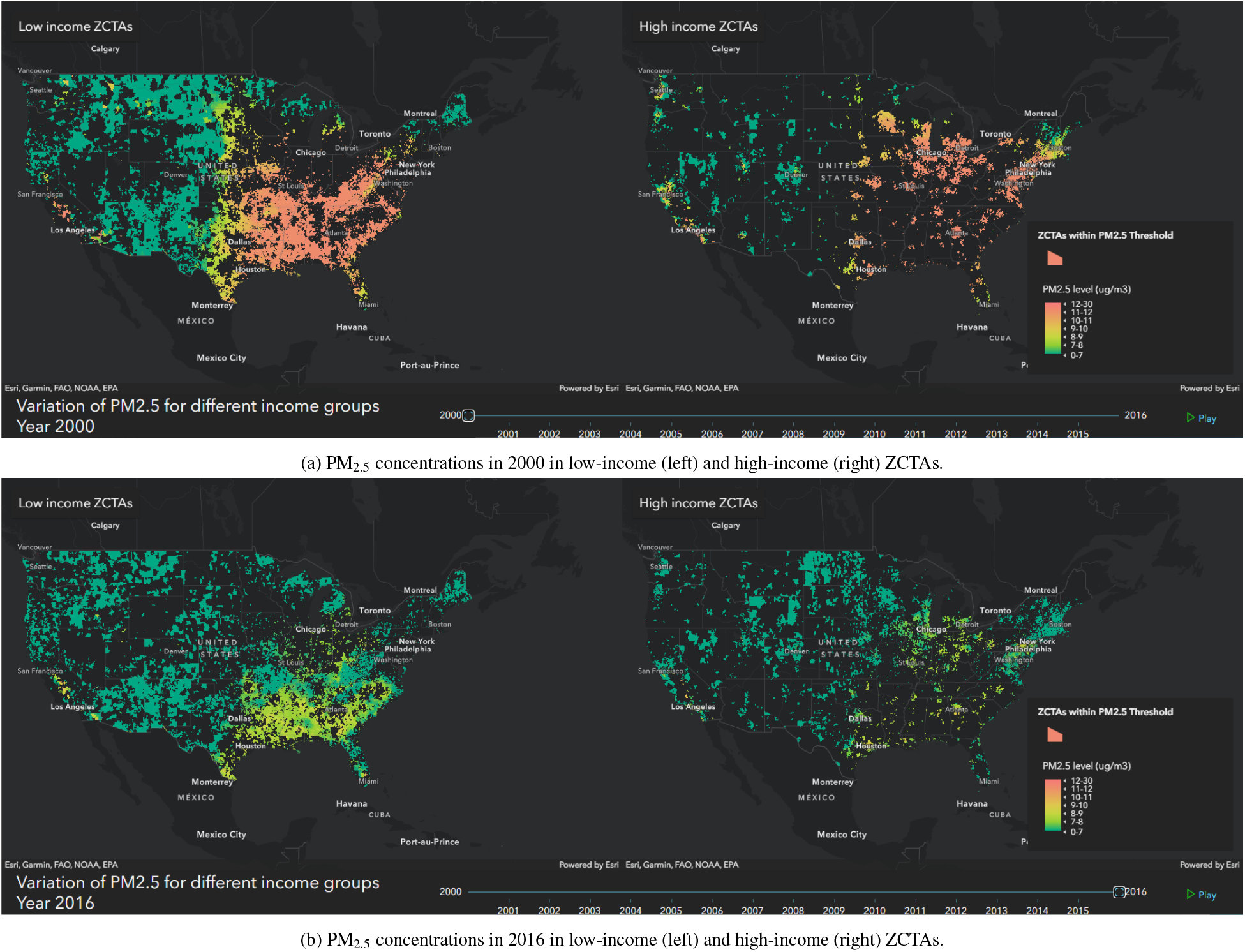
We assign all ZCTAs percentile ranks from 1 to 100 based on median household income and categorize them into ten income groups. We designate the lowest and highest three income groups as low-income and high-income respectively. The maps in panel (a) show PM_2.5_ levels for the year 2000 in low-income (left) and high-income (right) ZCTAs. Panel (b) shows the same information for the year 2016. Disparities in exposure to PM_2.5_ among the two groups are apparent and it can be visually seen that a larger decrease in PM_2.5_ concentrations from 2000 to 2016 occurred in high-income ZCTAs (Animation 3 in supplementary material).

## Racial and income inequalities in exposure to air pollution are increasing

We found that although air pollution in the US has decreased, inequality in breathing polluted air has increased among the different racial/ethnic and income groups. To visualize this finding, for each year and for each race, we ranked the US ZCTAs from the least dense to the most dense with respect to that race^3^ (Extended Data figure A.7a). For example, in figure 3a the dark blue region on the Black population map contains the ZCTAs with the highest ratio of Black population to total ZCTA population, and the light yellow region contains the ZCTAs with the lowest ratio of Black population to total ZCTA population. Similarly, the White population map in figure 3a contains dark blue and light yellow regions that correspond to high and low White population proportions respectively. In figure 3a, we only show for the Black and White populations the ZCTAs with a PM_2.5_ concentration higher than 8 *μg/m*^3^ for the year 2010 (i.e. high pollution ZCTAs). This figure reveals that almost half of these high pollution ZCTAs are where the Black population is concentrated (southern part of the map as indicated by the dark blue region on the Black population US map), and the other half is where the White population is concentrated (northern part of the map as indicated by the dark blue region on the White population US map). However, reexamining the high pollution ZCTAs for 2016 (figure 3b) shows that only those with concentrated Black populations remain above the PM_2.5_ threshold (majority of the Black population map is dark blue and that of the White population map is light yellow). In summary, air pollution reduction regulations between 2010 and 2016 were enforced and attained in the areas dominated by the White populations whereas the Black populations are still living with high PM_2.5_ levels; this clearly indicates an increase in disparities to air pollution exposure among the White and Black populations as will be numerically shown later. Additionally, we extend figure 3 to include the Hispanic and Asian populations and present the results in Extended Data figures A.7b and A.7c. We also repeat the same visualization for thresholds different than 8 *μg/m*^3^ and present them in Animation 4 in Supplementary Material, which shows the distribution of the different racial/ethnic communities across multiple PM_2.5_ concentrations for 2010 and 2016.

**Figure 3:**
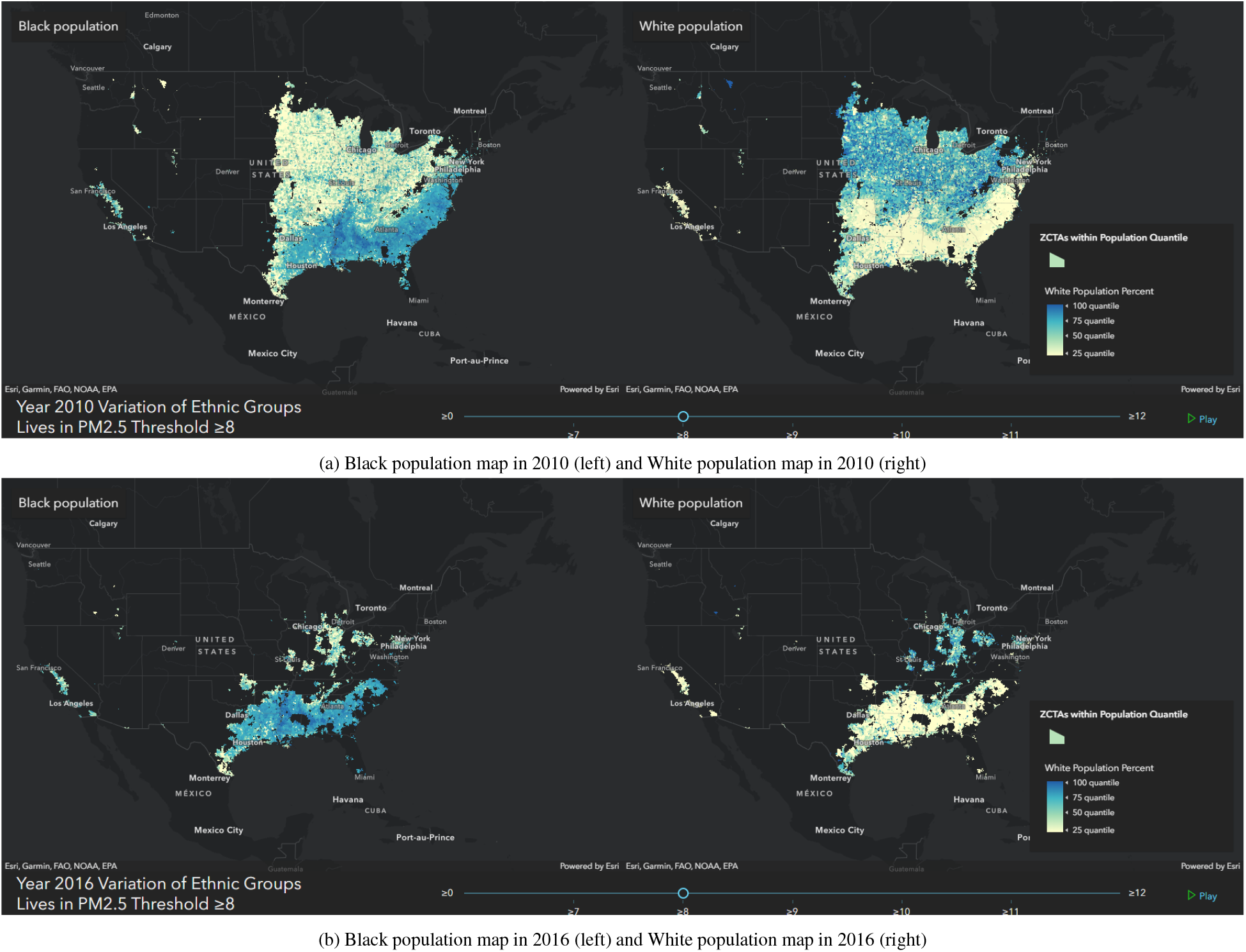
The maps only show US ZCTAs with PM_2.5_ levels above 8 *μg/m*^3^ in (a) 2010 and (b) 2016. We first focus on the maps on the left of the figure which correspond to the Black population. A color spectrum from light-yellow to dark-blue is used to color-code the ZCTAs, where the dark-blue and light-yellow colors correspond to ZCTAs with the largest and smallest Black population percentages of the total ZCTA population respectively, or equivalently where the Black population is most and least present respectively. The Black population map in panel (a) reveals that almost half of the ZCTAs with PM_2.5_ levels above 8 *μg/m*^3^ in 2010 correspond to those where the Black population most lives (almost half of the map is dark-blue). However in 2016, ZCTAs that remained above 8 *μg/m*^3^ are only those that are dominated by the Black population (Black population map in panel b). Similarly, the maps on the right of the figure correspond to the White population, where dark-blue and light-yellow colors corresponds to ZCTAs with the largest and smallest White population respectively. Contrary to the previous finding, ZCTAs that remained above 8 *μg/m*^3^ are only those where the White population is least present (White population map in panel b). This reveals that reduction in air pollution from 2010 to 2016 occurred in ZCTAs where the Black and White populations are least and most present respectively. Animation 4 shows the distribution of the different racial/ethnic communities across multiple ranges of PM_2.5_ concentrations in 2010 and 2016.

Furthermore, we quantify such variation in disparities to air pollution exposure among racial/ethnic (and income) groups using the coefficient of variation *CoV* (please see Methods) and summarize our findings in figure 4. For example, interpreting the figures for 2010 and 2016 (shown in figures 3a and 3b), we find that 75% of the population was exposed to PM_2.5_ levels higher than 8 *μg/m*^3^ in 2010, whereas only 36% in 2016 (solid blue line). However, the *CoV* shows that the variation in air pollution exposure among racial/ethnic groups relative to its mean increased from 0.17 in 2010 to 0.27 in 2016, which shows that exposure disparities among racial/ethnic groups have increased by a factor of 1.58. This is in agreement with figure 3. It is interesting to note that inequality in exposure to PM_2.5_ levels above 8 *μg/m*^3^ remained constant between 2010 and 2014, and started to increase in 2015. Figure 4a shows the analysis for a threshold *T* = 10 *μg/m*^3^, which reveals greater inequalities. The analysis is also applied for the case of the ten income groups instead of the racial/ethnic groups (figure 4b). We observe that inequality in exposure to PM_2.5_ levels above 10 *μg/m*^3^ across income groups decreased from 2010 to 2013 and then started to increase. Further, the level of inequality among income groups as measured by *CoV* is smaller than that observed among racial/ethnic groups.

**Figure 4:**
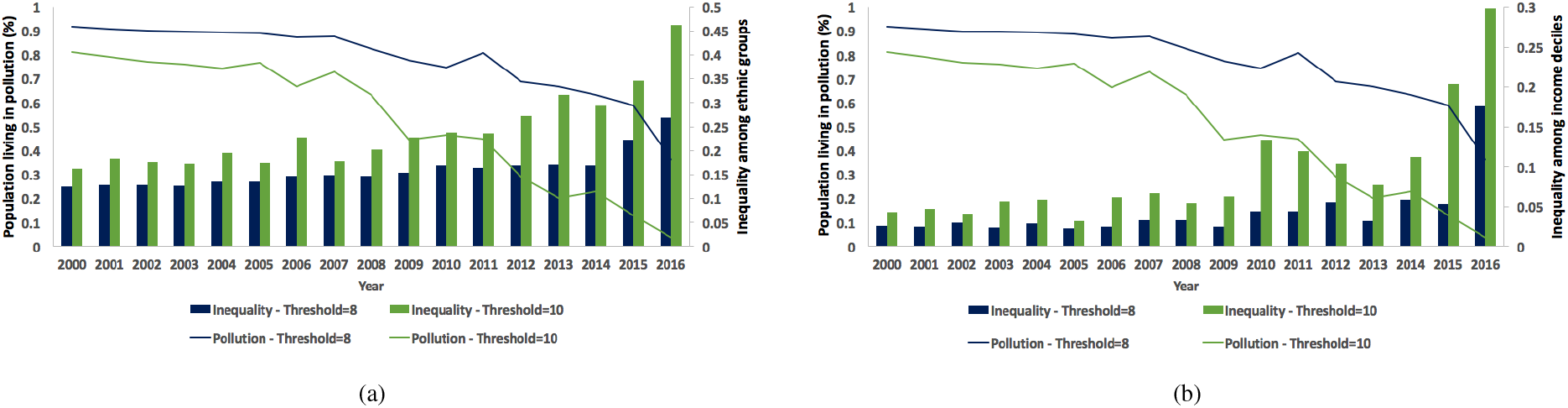
Inequality in exposure (as measured by *CoV*) to PM_2.5_ concentrations above thresholds of 8 and 10 *μg/m*^3^ for 2000 to 2016 among (a) racial/ethnic groups (Black, White, Hispanic, Asian, and Native American) (b) and income groups (income deciles). The percentage of the US population living above the thresholds of 8 and 10 *μg/m*^3^ is also shown. The trend reveals that the decrease in air pollution has been accompanied by an increase in the disparities in exposure to air pollution among communities.

In addition to using the easily interpretable *CoV*, we repeated the inequality analysis of figure 4 using the Atkinson index, an alternative inequality metric used in the literature [16, 20, 21]. These findings are located in Extended Data figure A.8 and are similar to those of figure 4 which confirms the increase in disparities in exposure to air pollution among racial/ethnic groups and among income groups.

## Discussion

We built a dataset that includes all US ZCTAs^4^ for 2000 to 2016 and contains demographic (median household income, racial/ethnic distribution, etc.) and pollution data (PM_2.5_ concentrations) and investigated temporal patterns in PM_2.5_ levels and in air pollution exposure of racial/ethnic and income groups. PM_2.5_ concentrations decreased drastically from 2000 to 2016, where the population-weighted average of PM_2.5_ has decreased by 43% from the year 2000 (13 *μg/m*^3^) to 2016 (7.3 *μg/m*^3^). Further, it was evident that air pollution exposure disparities exist among both racial/ethnic and income groups. When investigating the air pollution exposure of the different racial/ethnic communities, we found that the Black population is consistently exposed to higher levels of PM_2.5_ on average. In 2016, the average PM_2.5_ concentration for the Black population was 11.6% higher than that of the White population and 28.6% higher than that of the Native American population. The Native American population was consistently exposed to the lowest levels of air pollution from 2000 to 2016 (Extended Data figure A.4a). Additionally, we found that as the Black population in ZCTAs increased, the average PM_2.5_ concentration increased; a steep increase was observed in ZCTAs where the Black population exceeded 85% of the population (Extended Data figure A.4b). Contrary to trends of the Black populations, the average PM_2.5_ concentration decreased as ZCTAs became more populated with the White and Native American populations (Extended Data figure A.4b). The analysis also showed that the low-income group consistently suffers from worse air conditions compared to higher-income groups. In 2016, the average low-income population PM_2.5_ concentration was 6.9% higher than that of the high-income population (Extended Data figure A.5). Of note, exposure disparities in racial/ethnic and income groups are related to geographical distribution. For example, the industrial nature of southern US may result in less strict pollution regulations and is more polluted. At the same time, southern states have higher Black and low income populations.

The most pertinent and concerning finding of our study is that even though air pollution has decreased, exposure disparities among both racial/ethnic groups and income groups (as measured by the coefficient of variation and Atkinson index) have increased. Indeed, figure 3 shows that air pollution reduction strategies from 2010 and 2016 mainly resulted in air pollution reduction in areas where the White population is most concentrated.

The results of increasing inequalities in air pollution exposure are timely and concerning at a time where the US is facing crises of racism, and health disparities in COVID-19 health outcomes. There is an emerging research area in the US and around the world that provides preliminary evidence that long-term air pollution exposure increases susceptibility to COVID-19 [22]. Furthermore, it has been documented that racial minorities have higher COVID-19 death rates [23, 24]. The results of this paper show that the EPA must not only act to decrease nationwide PM_2.5_ concentration levels on average but must also devise air pollution reduction strategies to reduce pollution exposure of minority and low-income populations to address environmental injustice.

## Data Availability

Data will be made accessible upon manuscript acceptance in a peer reviewed journal through the publishing journal.

## Supplementary Material

Supplementary material is available upon request from corresponding author.

## Data Availability

Data will be made accessible through the publishing journal upon manuscript acceptance.

## Code Availability

The code will be made accessible through the publishing journal upon manuscript acceptance.

Acknowledgements

The authors would like to thank Joel D. Schwartz for providing the air pollution data, Benjamin Sabbath for cleaning and preparing the datasets, and Lauren Bennett for comments and discussions. This work was supported financially by grants from the Health Effects Institute (4953-RFA14-3/16-4), National Institute of Health (DP2MD012722), National Institute of Health and Yale University (R01MD012769), National Institute of Health and National Institute of Environmental Health Sciences (R01 ES028033) and the Environmental Protection Agency (83587201-0).

## Author contributions

AJ, SV and FD contributed to the study design. AJ led the research with supervision from FD. XZ, JL and THL prepared the map animations. AJ drafted the manuscript with support from XZ, JL, THL, SV and FD. All authors read and approved the final manuscript for submission.

## Methods

Our dataset includes all US zip code tabulation areas^5^ (ZCTAs) for 2000 to 2016 (N=32047). For each ZCTA, we obtained demographic and socioeconomic variables from the US Census Bureau when available and used interpolation techniques (moving average) to determine those of the missing years. Variables of interest comprised median household income, proportions of Native Americans, Asian, White, Black, and Hispanic residents, and population density. For each year, we assigned all ZCTAs percentile ranks from 1 to 100 based on median household income and categorized them into ten income groups. Throughout the paper we use low-income and high-income to label the lowest three and highest three income groups respectively.

We also used previously in-house developed prediction models to estimate PM_2.5_ concentration levels on 1 km by 1 km nationwide grids [25, 26]. These use a validated ensemble-based model, which integrates three machine learning (ML) algorithms including gradient boosting, neural network, and random forest. These ML algorithms used more than 100 predictor variables from satellite data, land-use terms, meteorological variables, and chemical transport model predictions. The ensemble-based model was calibrated using daily PM_2.5_ concentrations measured at 2,156 US EPA monitoring sites, with an average cross-validated *R*^2^ of 0.86 and an unbiased slope, indicating excellent model performance. For each ZCTA we calculated the daily average PM_2.5_ based on all covered 1 km^2^ grid cells and then computed annual averages. The annual averages of PM_2.5_ levels for each ZCTA were included in the dataset. We built one dataset by combining the demographic and PM_2.5_ variables across all US ZCTAs for 2000 to 2016. Our dataset analysis reveals time patterns in air pollution across the US and inequalities in exposure to air pollution among racial/ethnic and income groups. Dynamic animations are used to communicate our findings along with plots that summarize and clarify the information embedded in our visualizations.

We first defined a group population-weighted PM_2.5_ concentration, where a group can be an income group such as the first decile, or an ethnic group such as the Hispanic population. In the case of racial/ethnic groups, the population-weighted PM_2.5_ concentration in racial/ethnic group *k* is given by:

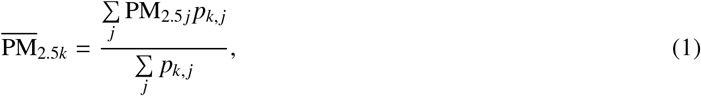

where summation occurs over all ZCTAs. *p*_*k, j*_ is the number of people in racial group *k* living in the ZCTA *j*, and PM_2.5 *j*_ is the PM_2.5_ level in the ZCTA *j*. In the case of income groups, the population-weighted PM_2.5_ concentration of income group *i* is:

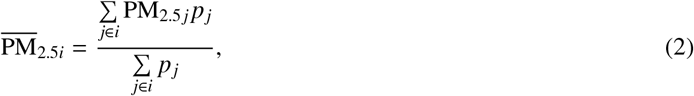

where summation occurs only over ZCTAs *j* belonging to the income group *i. p*_*j*_ is the total population of ZCTA *j*, and PM_2.5 *j*_ is the PM_2.5_ level in ZCTA *j*.

Additionally, we defined a second PM_2.5_-related variable (*q*) and use it to quantify the level of inequality in exposure to PM_2.5_ concentrations among the different racial/ethnic or income groups. The variable *q* is defined as the percentage of a population exposed to PM_2.5_ levels above a certain PM_2.5_ threshold *T*. We can calculate *q* for specific population subgroups. For example, we can compute the percentage of the population in the highest income group that is exposed at PM_2.5_ levels above *T* = 12 *μg/m*^3^, or the percentage of a racial/ethnic group, such as Native Americans, exposed to PM_2.5_ levels above *T* = 8 *μg/m*^3^.

To measure the degree of inequality across racial/ethnic (or income) groups in exposure to PM_2.5_ concentrations above *T* for a specific year, we first compute *q* for every racial/ethnic (or income) group. We then compute the coefficient of variation (*CoV*) defined as:

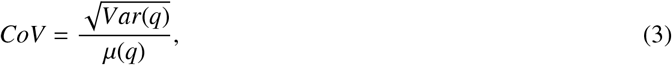

where *Var* is the variance of *q* and *μ* is the mean of *q. CoV* measures the variability of a series of numbers independent of the data magnitude, so it captures the variation in *q* among racial/ethnic (or income) groups in a given year relative to the mean pollution levels during that year.

For example, consider three years *Y*_1_, *Y*_2_ and *Y*_3_, where the percentages of five racial/ethnic groups being exposed to PM_2.5_ levels above a threshold *T* are, respectively:

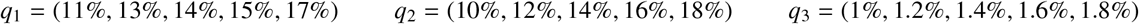

From *Y*_1_ to *Y*_2_, the coefficient of variation increases from *CoV*_1_ = 0.143 to *CoV*_2_ = 0.202, which indicates that the variation in exposure to air pollution relative to the mean, and equivalently disparities among the racial/ethnic groups, increased by a factor of 1.4. On the other hand, although the pollution levels decreased drastically between *Y*_2_ and *Y*_3_ as can be seen by the different orders of magnitude of *q*_2_ and *q*_3_, the coefficient of variation is unchanged (*CoV*_3_ = 0.202) indicating that the inequalities in exposure to air pollution among the racial/ethnic groups is the same between *Y*_2_ and *Y*_3_. These examples highlight the power of using *CoV* to capture relative variation in the data independently of its magnitude. This is very important for our application because the level of pollution changes considerably over the years.

The outlined procedure of quantifying inequality through *CoV* can be applied for any PM_2.5_ threshold *T* and can be repeated for all years to track the evolution of inequality in exposure to air pollution among the different racial/ethnic (or income) groups.

## Appendices

### A. Extended Data Figures

**Figure A.1:**
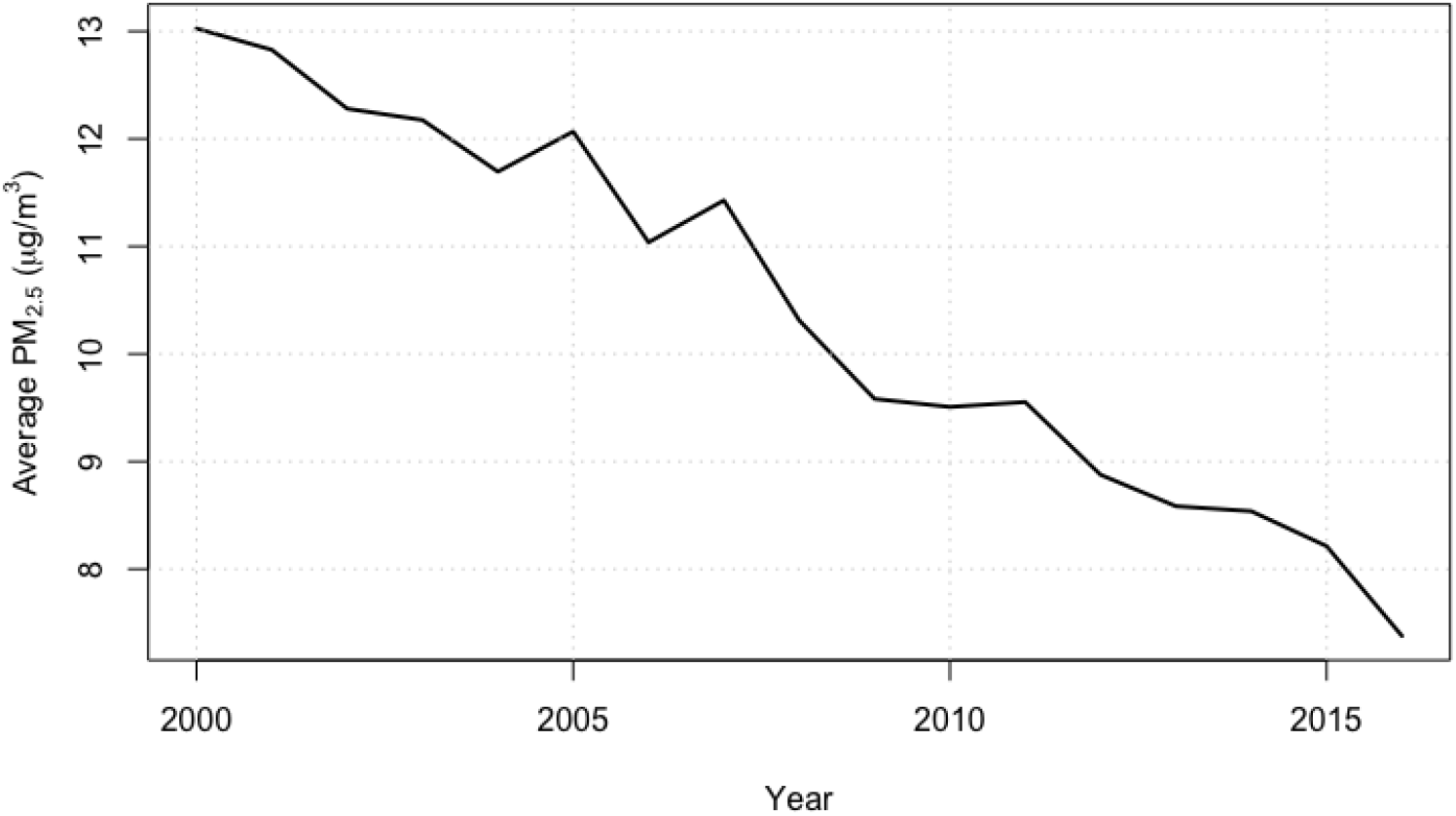
The national average PM_2.5_ level has been decreasing in the US since 2000.

**Figure A.2:**
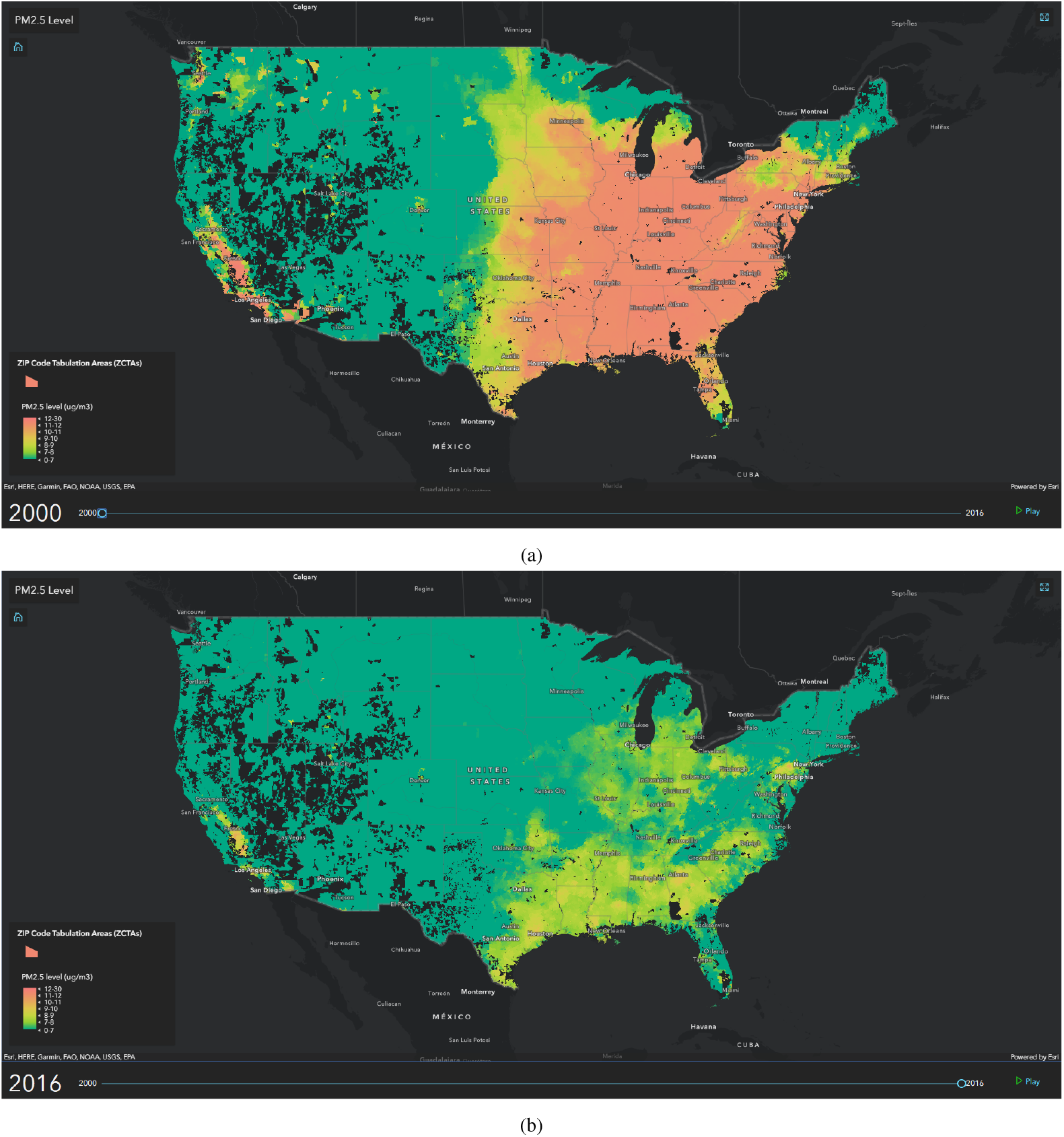
Distribution of PM_2.5_ concentration levels in (a) 2000 and (b) 2016. We also include an animation that shows the distribution of PM_2.5_ concentration levels in the US from 2000 to 2016. (Animation 1)

**Figure A.3:**
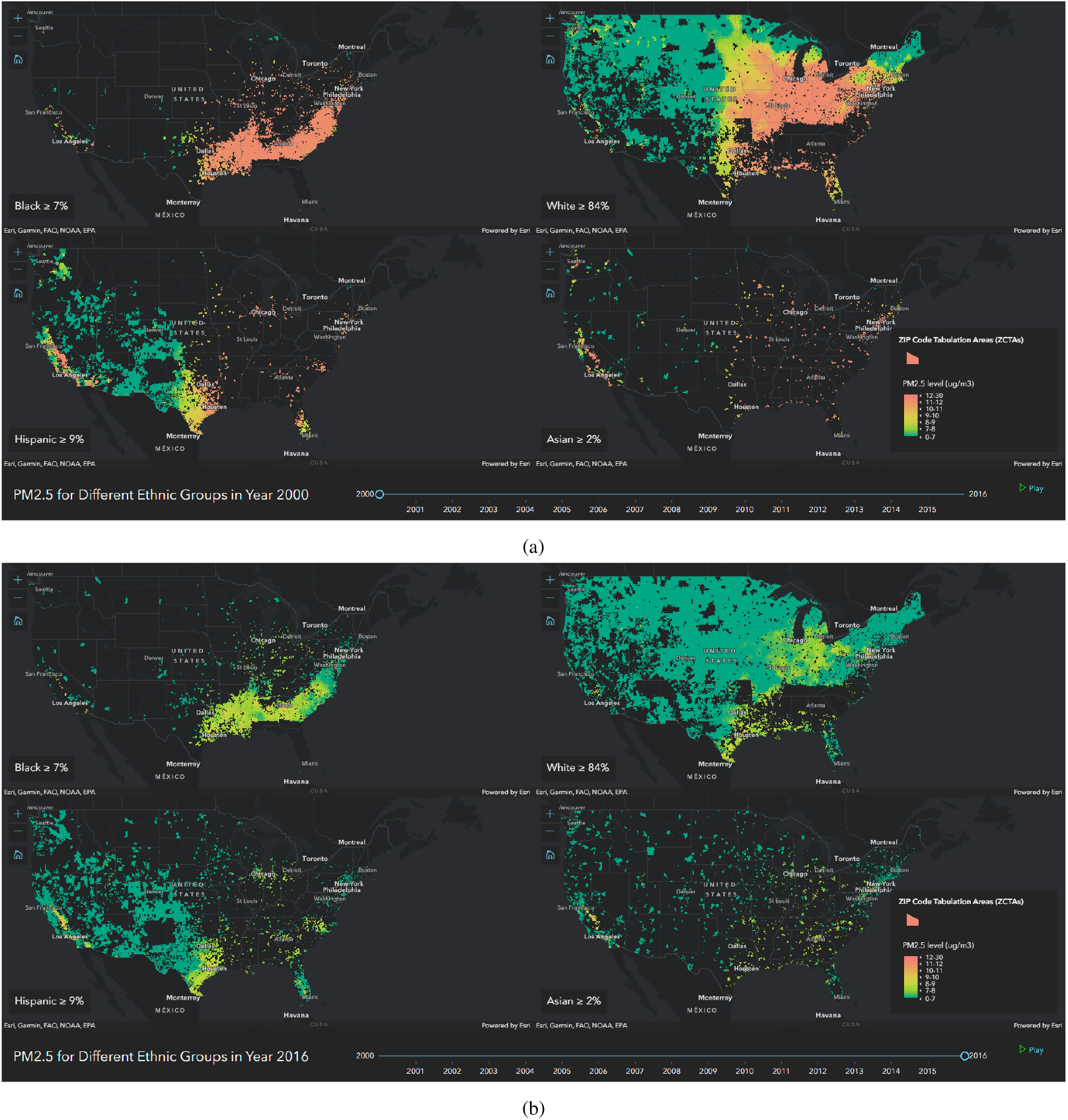
Distribution of PM_2.5_ across four different maps each corresponding to a race/ethnicity for (a) 2000 and (b) 2016. (Animation 2)

**Figure A.4:**
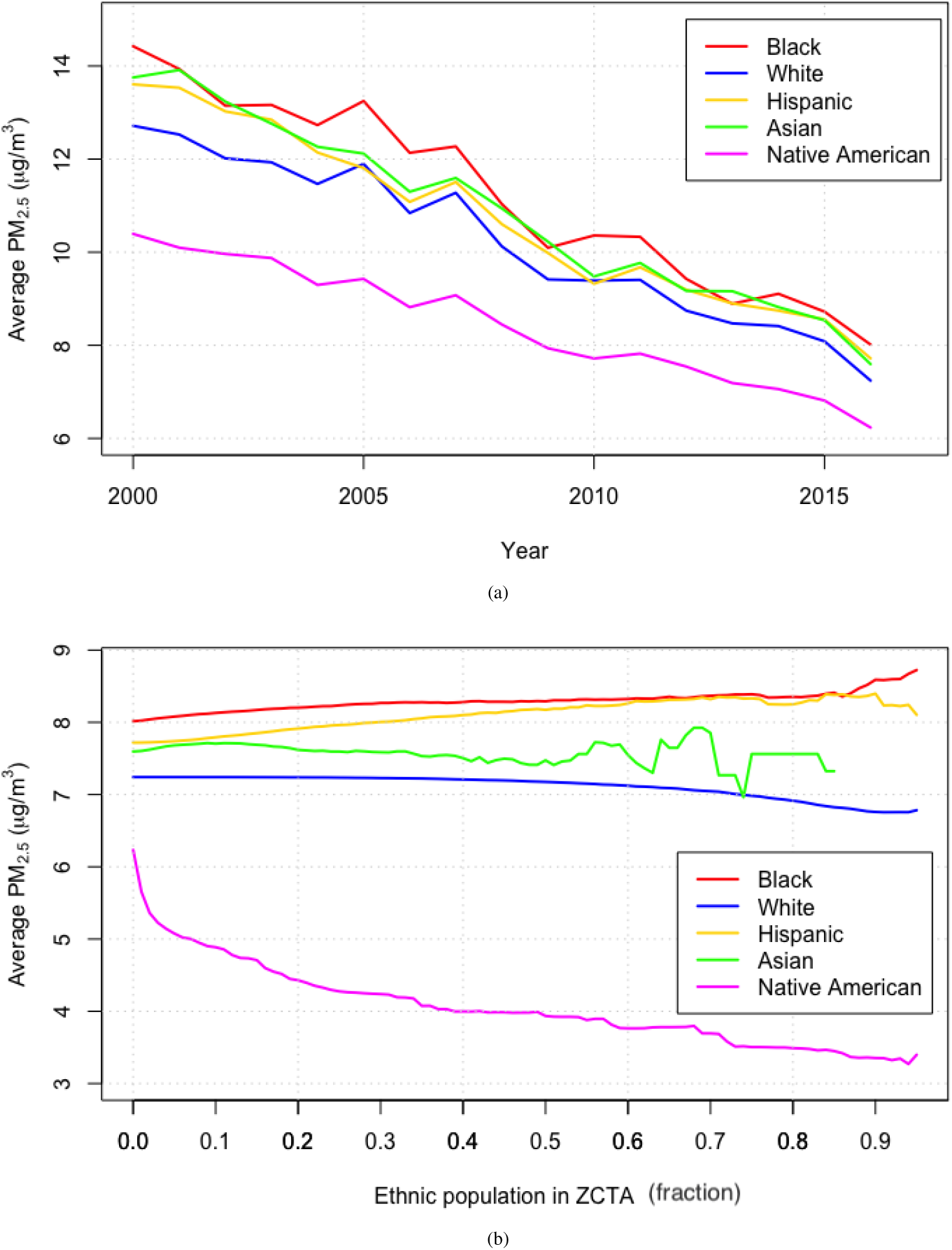
(a) The population-weighted PM_2.5_ average concentration across the different racial/ethnic communities for 2000 to 2016. The PM_2.5_ concentration across the racial/ethnic communities demonstrates that Black and Native American populations live in the most and least polluted areas respectively. (b) The population-weighted PM_2.5_ average concentration across racial/ethnic communities as a function of ZCTA racial/ethnic population (%) for 2016. When the racial/ethnic population % is equal to 0.2, the red curve includes every ZCTA where the Black population is 20% or more, and the blue curve includes every ZCTA where the White population is 20% or more. As ZCTA’s Black and Hispanic populations increase, the PM_2.5_ concentration levels increase. The opposite effect is seen for the White and Native American communities.

**Figure A.5:**
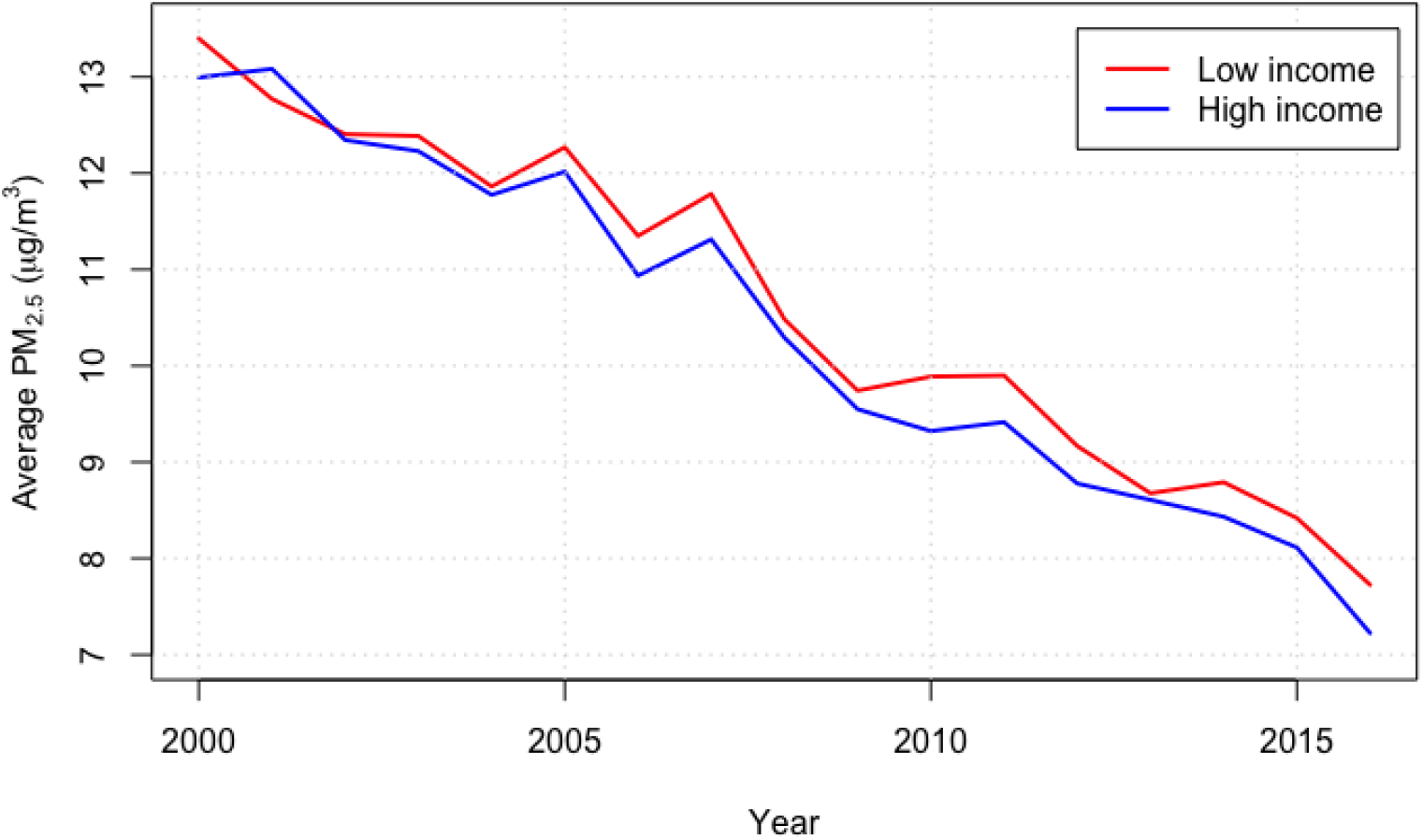
The PM_2.5_ concentration across the income groups reveals that the low-income group is located in the most polluted areas.

**Figure A.6:**
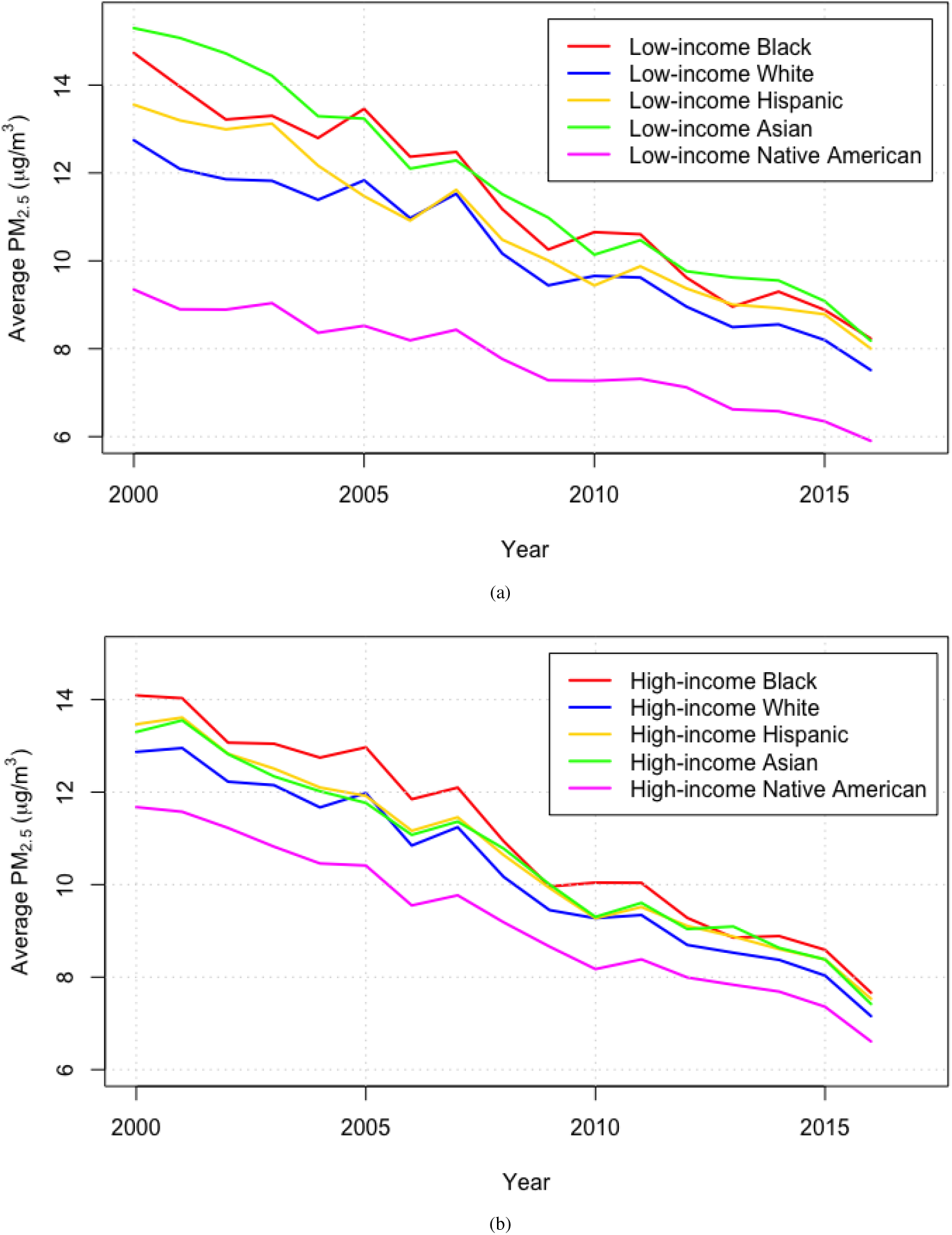
(a) The population-weighted PM_2.5_ average concentration across the different racial/ethnic communities for 2000 to 2016 that are in the low-income group. A similar trend to that shown in Extended Data figure A.4a is seen except for the Asian population. (b) The population-weighted PM_2.5_ average concentration across the different racial/ethnic communities for 2000 to 2016 that are in the high-income group. A similar trend is seen in Extended Data figure A.4a. It is interesting to note that for the Native American population, low-income groups are exposed to lower concentrations of PM_2.5_ as compared to the high-income groups. This may be tied to the rurality vs. urbanicity of the ZCTAs, which was not included in our analysis.

**Figure A.7:**
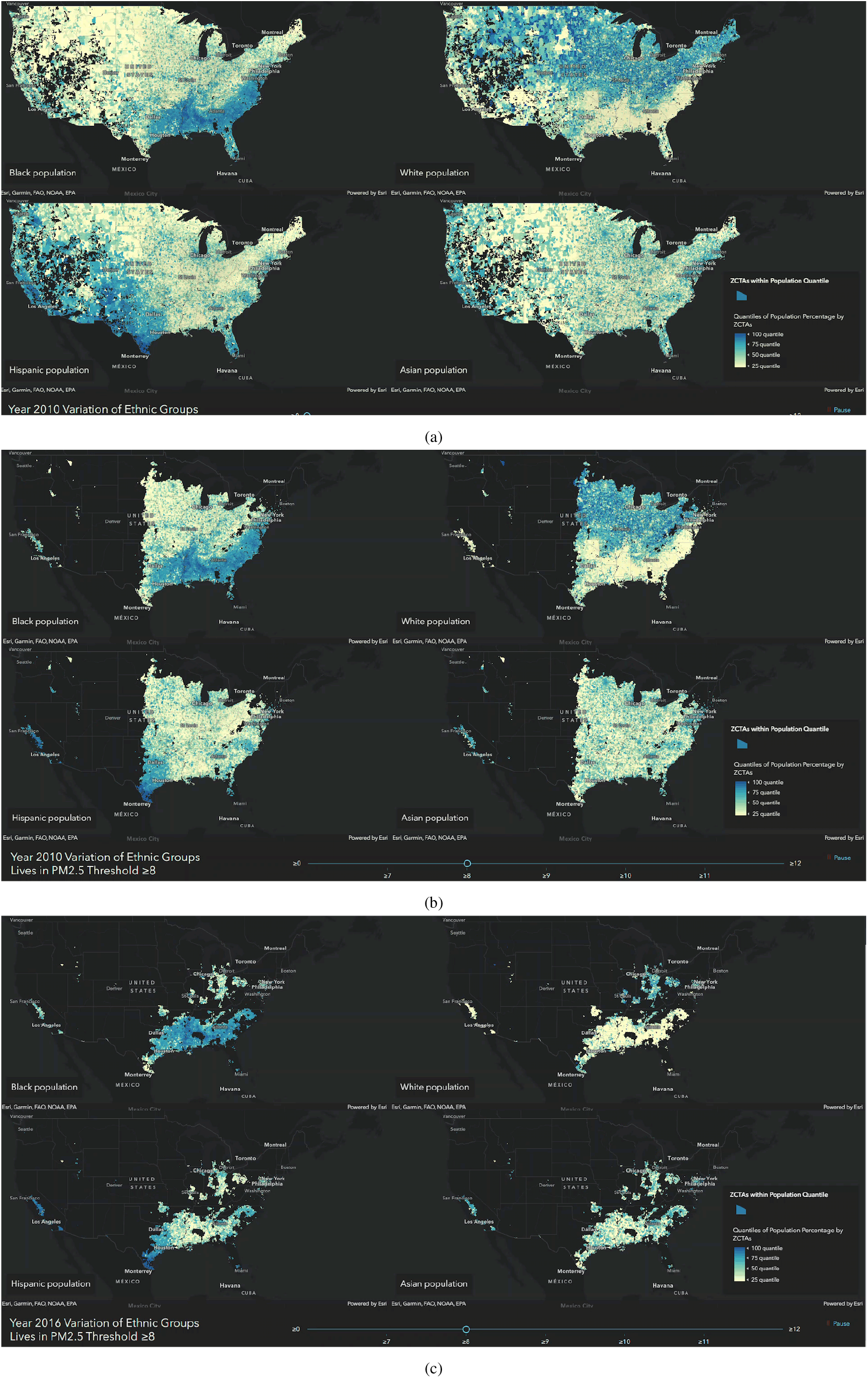
The distribution of the racial/ethnic populations above a PM_2.5_ threshold of 8 *μg/m*^3^ for 2010 and 2016. (a) US ZCTAs for each race/ethnicity are ranked based on the ratio of the race/ethnicity population to the total ZCTA population. Dark blue indicates fractions close to 1 (ZCTAs where the corresponding race/ethnicity most lives), and light yellow indicates fractions close to 0 (ZCTAs where the corresponding race/ethnicity least lives) (b) US ZCTAs above 8 *μg/m*^3^ in 2010. (c) US1Z6CTAs above 8 *μg/m*^3^ in 2016. Animation 4 shows the distribution of the different racial/ethnic communities across multiple ranges of PM_2.5_ concentrations for 2010 and 2016.

**Figure A.8:**
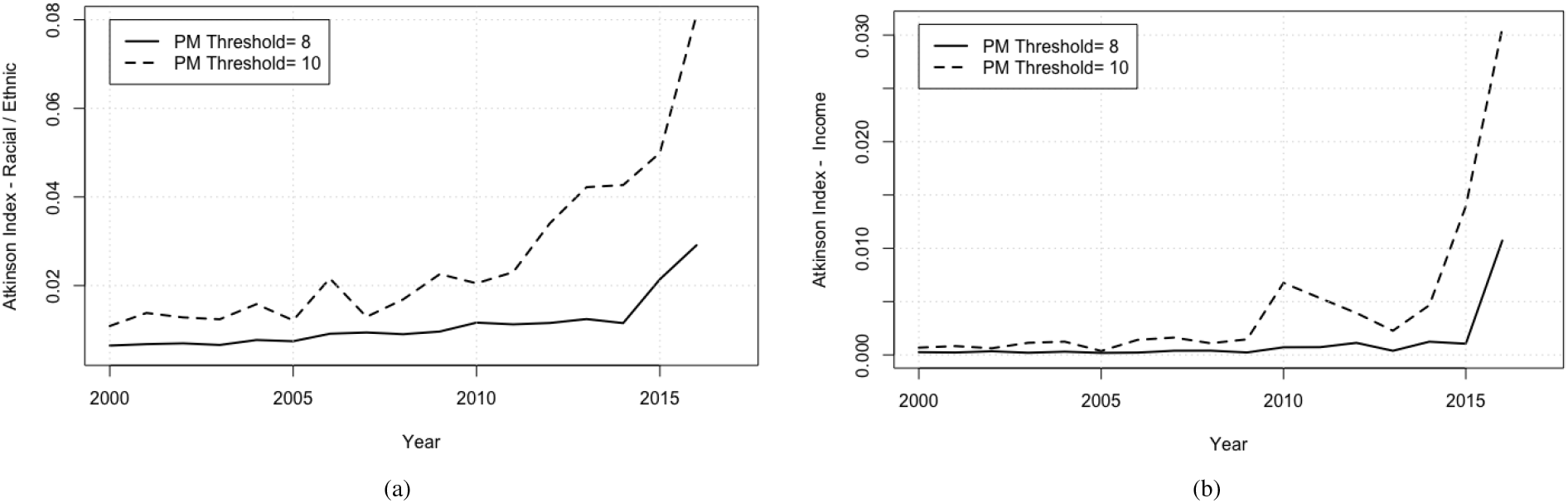
Inequality measured by the Atkinson index in exposure to PM_2.5_ concentrations above thresholds of 8 and 10 *μg/m*^3^ for 2000 to 2016 among (a) racial/ethnic groups (Black, White, Hispanic, Asian, and Native American) (b) and income groups (income deciles). The trend is similar to the one measured by *CoV* in figure 4, disparities in air pollution exposure among racial/ethnic groups and income groups are increasing. Further, disparities among racial/ethnic groups are higher than those among income groups, and disparities above a PM_2.5_ threshold of 10 *μg/m*^3^ are higher than above 8 *μg/m*^3^. The Atkinson index was computed using the inequality package “ineq” in the R software. The input is the proportion of the racial/ethnic (or income) groups living above the set PM_2.5_ threshold. We set the Atkinson aversion parameter *ϵ* = 0.75 [16].

## Footnotes

1 Native American was not included in the animation as regions of presence are very small and difficult to visualize. It was included in the main analysis.

2 In the case of the White population for example, we use the White population fraction of the ZCTA population to compute the average White population fraction across all ZCTAs (≈84.2%). We then set the margin at 84% and only show ZCTAs with a White population fraction exceeding this margin. The margin for the Black population was computed similarly (7.6%) and set at 7%.

3 Ranking the ZCTAs for a particular race is done by using the population fraction of the race in every ZCTA to split them into 100 quantiles by using percentiles.

4 Demographic data for multiple ZCTAs (2.7%) were missing. As a result, these geographic areas were not included in the analysis or animations.

5 Demographic data for multiple ZCTAs (2.7%) were missing. As a result, these geographic areas were not included in the analysis or animations.

